# Covid19 Surveillance in Peru on April using Text Mining

**DOI:** 10.1101/2020.05.24.20112193

**Authors:** Josimar Edinson Chire Saire, Jimy Frank Oblitas Cruz

## Abstract

The present outbreak as consequence by coronavirus covid19 has generated an big impact over the world. South American countries had their own limitations, challengues and pandemic has highlighted what needs to improve. Peru is a country with good start with quarantine, social distancing policies but the policies was not enough during the weeks. So, the analysis over April is performed through infoveillance using posts from different cities to analyze what population was living or worried during this month. Results presents a high concern about international context, and national situation, besides Economy and Politics are issues to solve. By constrast, Religion and Transport are not very important for peruvian citizens.

## I. Introduction

Public health vigilance is the practice of public health agencies that collect, manage, analyze and interpret data systematically and continuously, and spread such data to programs facilitating the measures in public health [1]. In this field, many ways of public health analysis appear, among them, infodemiology, an emerging area of research studying the relationship between information technology and consumer health, as well as the tools of infometrics and web analysis whose final objective is to inform and collaborate with public health and public policies [2]. Along with this, it is necessary to find determining disease outbreaks in advance in order to reduce their impact on the populations. The supposed advantage of getting information provided by automated systems falls short facing the impossibility of accessing data in real time, as well as inter-operational fragmented systems, which leads to the transfer and processing of longer data [3]. This kind of technology has been used for diseases, such as Whooping cough [4], flu [5], and immunosuppressive diseases [6], among others. Currently, we are facing coronavirus disease (COVID-19) which is a viral infection highly pathogenic caused by SARS-CoV-2. Currently, it is already causing global concern on health [7]. Officially declared as a global pandemic by the World Health Organization (WHO) on March 11, 2020, COVID-19 outbreak (Coronavirus 19 disease) has evolved at an unprecedented rate [8]. In order to help public health and to make better decisions regarding Public Health and to help with their monitoring, Twitter has demonstrated to be an important information source related to health on the Internet, due to the volume of information shared by citizens and official sources. Twitter provides researchers an information source on public health, in real time and globally. Thus, it could be very important for public health research [9]. Within the context of COVID 19, users from all over the world may use it to identify quickly the main thoughts, attitudes, feelings and matters in their minds regarding this pandemic. This may help those in charge to make policies, health professionals and public in general to identify the main problems that concern everybody and deal with them more properly [10]. This research is aimed to identify the main topics published by Twitter users related to the pandemic COVID19. Making the analysis of that information may help those in charge to make policies and healthcare organizations to assess the needs of interest groups and to deal with them properly.

The remainder of the paper follows. Section 2 presents related works regarding the retrieval infectious diseases information from social media. In section 3, the data collection methodology for extracting relevant information of Covid-19 from Twitter is presented. Section 4 describes experimental findings and a discussion related to the analysis. Finally, conclusions and future work are described in Section 5.

## II. Related work

Surveillance pretends to observe what happens over one population, region or city to support on Politics Decisions. One good advantage are cost and time because usually surveys are two components: collection and processing, both can spend many days, even months..

Sinnernberg [11] performs a study about Twitter as tool for Research on Public Health, is necessary to highlight researchers uses traditional databases for studies and Twitter can provide useful data from people. From 137 papers for the review, research fields as Public health (31), infectious disease (28).

Breland [12] express in Social Media people create content, exchange information and use this tool for communication. A four benefits from the use: a) disseminate Research on Public Health field, b) fight against misinformation, c) influence policies, d) aid Public Health Research and e) enhance professional development. And Yepes [13] can support the affirmation: Twitter is source from useful data for surveillance, considering relevant terms and geographical locations.

More applications using Twitter and Natural Language Processing are found: monitor H1N1 flu [14], Dengue in Brazil [15], covid19 symptomatology in Colombia [16], covid19 infoveillance in South America countries [17] and monitor City of Mexico [18]

Finally, Ear [19] found, Peruvian Internal Agencies have overlapping functions so this can limit collaboration, there is not enough technical capacity and resources outside the capital, Lima. Besides, cultural diversity and geographical issues can present challenges to fight agains one disease infection. Therefore, the use of a infoveillance tool based on Text Mining can provide a support to the goverment and public policies creation.

## III. Methodology

The process to analyze the situation in Peru, follows the next steps:

- Select the relevant terms related to covid19 pandemic
- Set the parameters to collect related posts
- Pre-processing
- Visualization

A. *Select relevant terms* The scope of the analysis is Peru, and this regions so considering news about Covid-19, the selected terms are:
  - ’coronavirus’,’covid19’, cuarentena, pandemia
B. *Build the Query and Collect Data* The collection process is through Twitter Search function, with the next parameters:
  - date: 01-04-2020 to 31-04-2020
  - terms: the chosen words mentioned in previous subsection
  - geolocalization: the capital of every state from Peru, see Fig. 1 1
  - language: Spanish
  - radius: around 50 km
C. *Preprocessing Data* This step is very important to take relevant words and this is the source to create graphics to help understanding the country.
  - Uppercase to lowercase
  - Eliminate alphanumeric symbols
  - Eliminate words with size less or equal than 3
D. *Visualization*

**Fig. 1.**
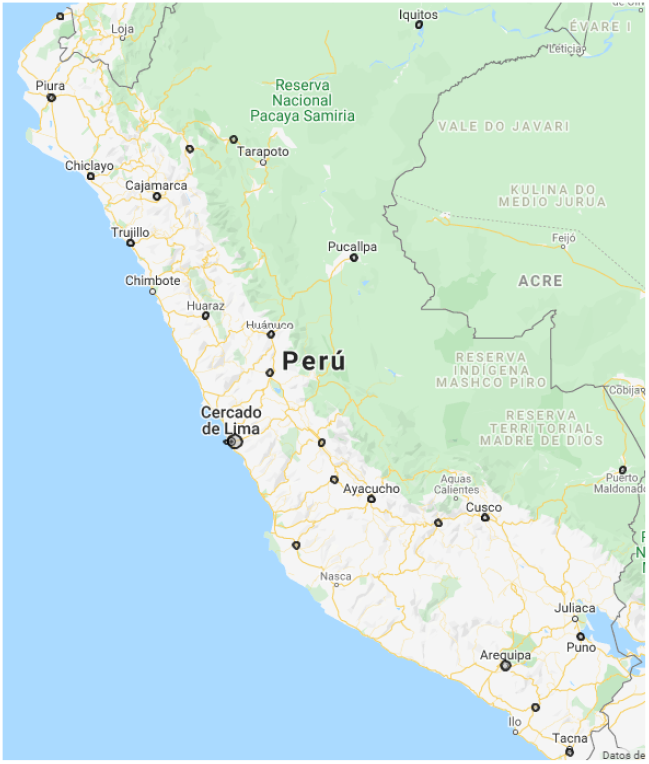
Geolocalization of Regions of Peru of Spanish Speakers

**Fig. 2.**
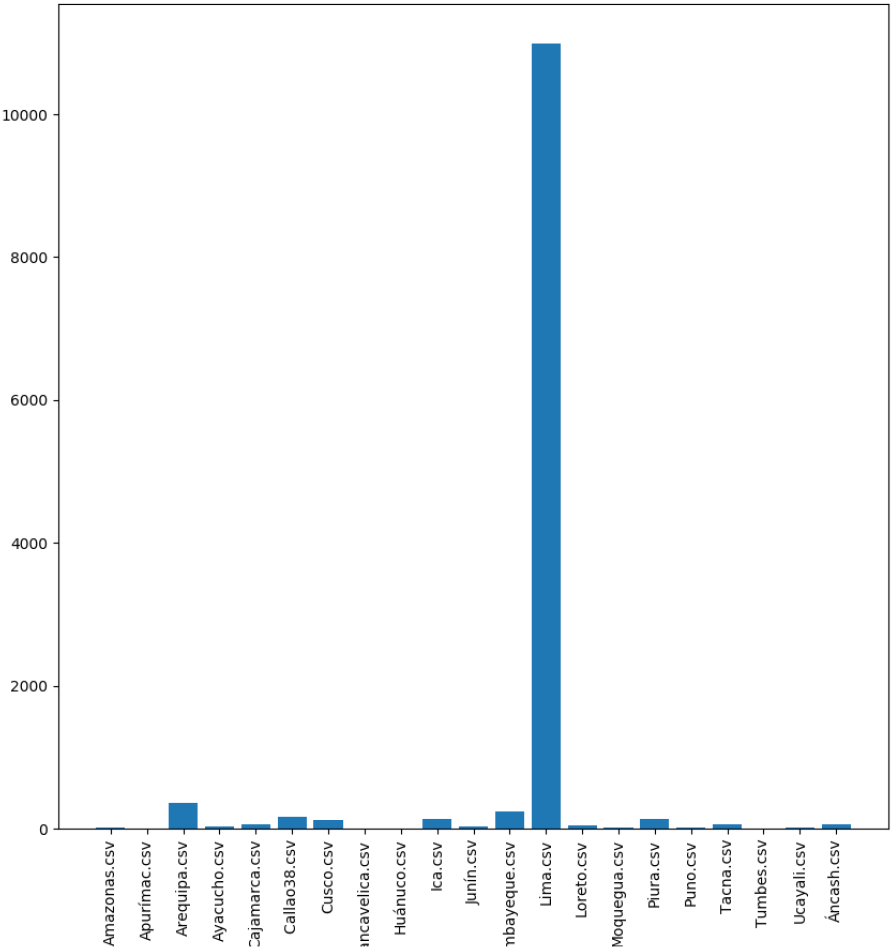
Data User Creation

## IV. Results

The next graphics presents the results of the experiments and answer some questions to understand the phenomenon of the pandemic over Perú country population.

Helping the visualisation from Monday to Sunday during the last two weeks, a cloud of words is presented in Fig. 3.

**Fig. 3.**
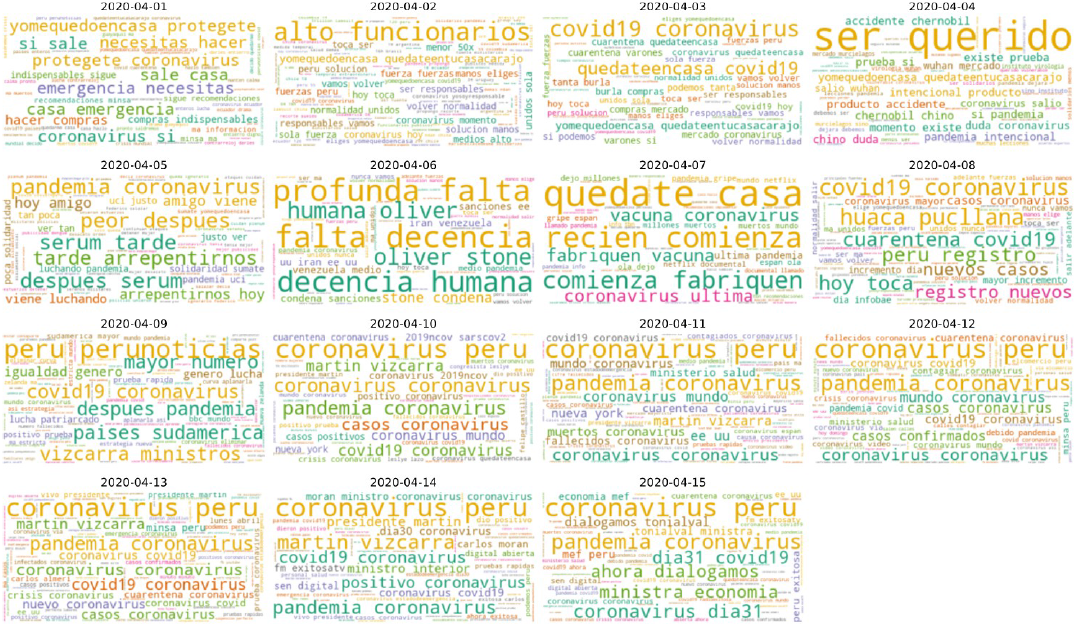
Cloud of Words: Lima

Analyzing Lima Fig.4, the one hundred of more frequent terms are related to cases of coronavirus and extracting a value between number of tweets and number of time for each team is natural to conclude the most important topic is related to health and covid19.

**Fig. 4.**
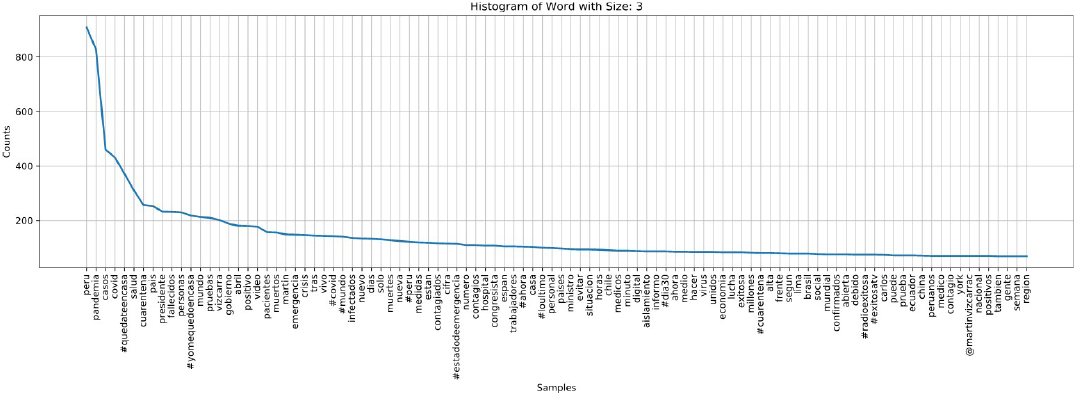
Word Histogram

According to the data analysis, it can be seen the regions including words related to “entertainment” issues such as, Lambayeque, La Libertad, Piura and Loreto, those which have the highest level of contagion in Peru. This issue is related to social and culture differences of the northern coast of the country. A similar case occurs when searching words related to “religious” issues, where regions such as, Cajamarca, Cuzco and Huanuco, include them due to their traditions, common in the zone. see Fig. 5, that Since the current situation has made the population to “abandon” some customs and to adopt other new ones, for many people it was hard to adopt these recommended measures firstly, so they has had to look for them by using social networks.

**Fig. 5.**
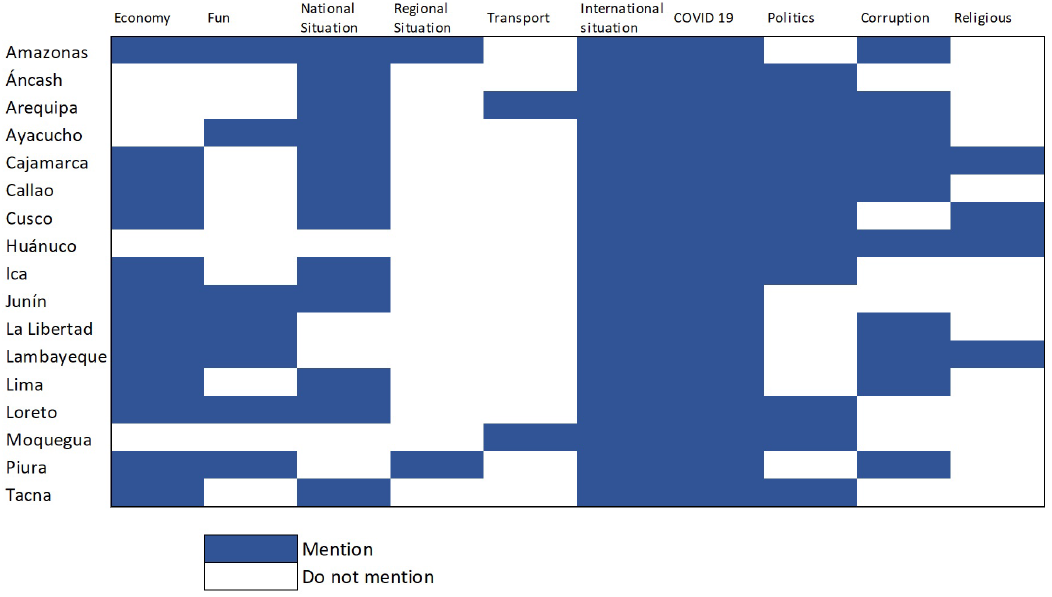
Caption

Another point to consider is that, besides the information on COVID 19, the international information related to the situation in other countries is present in every region. This kind of information is followed by domestic issues from the national situation. Publications referring to regional or local issues are scarcely present, see Fig. 6. This may be because, even though health is an aspect that causes society concern, regarding prevention, information coming from the national government is preferred.

**Fig. 6.**
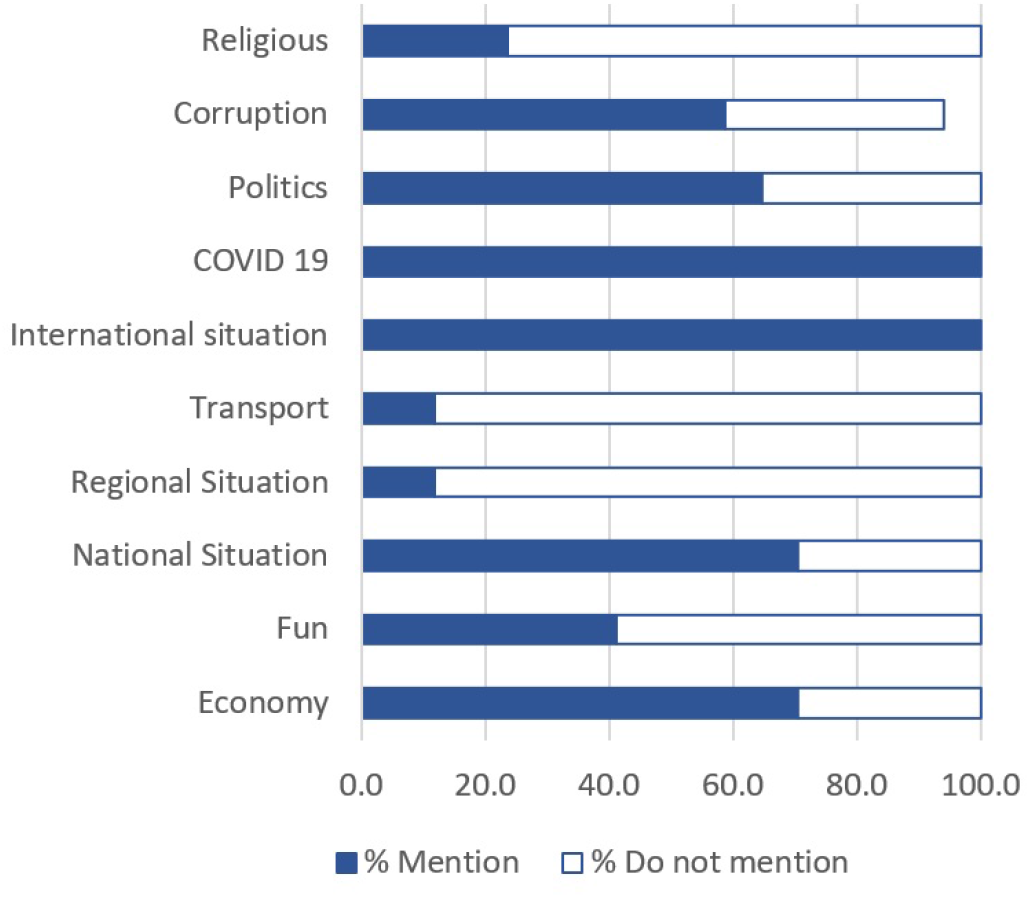
Caption

A similar scenario is present in all the regions, mass media is part of the top users and regular people is posting, so it is possible to know what they are thinking about covid19, see Fig. 7.

**Fig. 7.**
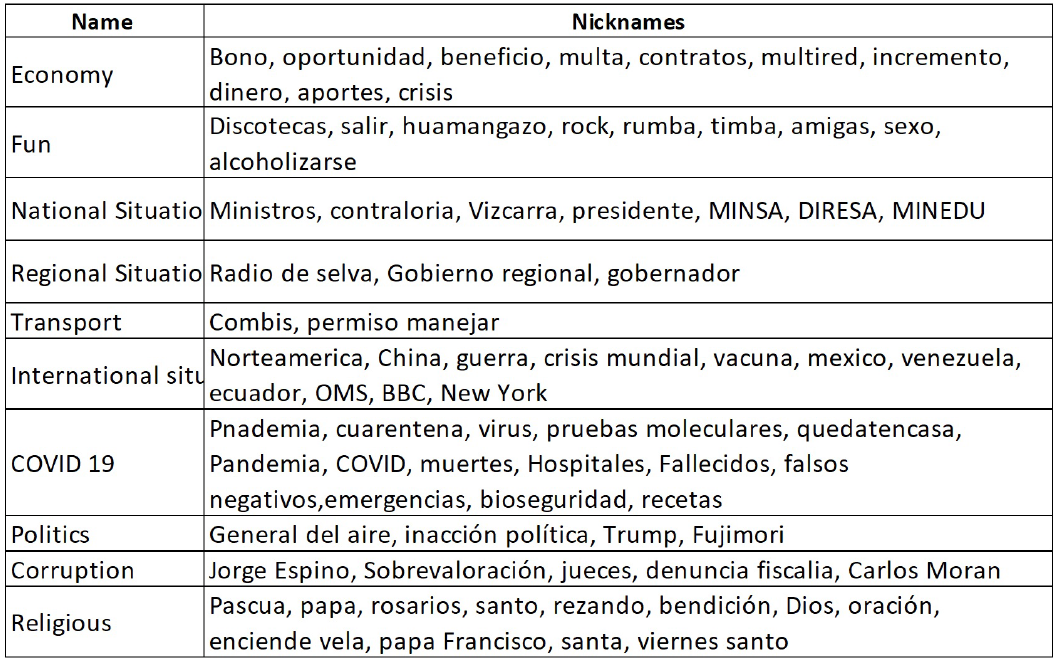
Caption

## V. Conclusions

The Social Network explored is useful to provide data for a exploratory analysis, to know what concerns can have citizens and map the issues per city so Public Policies can be more efficient and located. Peruvian Citizens have a high concern related to Covid19, international context and Economy, Politics from National Context and a minor worrying about Religion, Transport.

## Data Availability

Data will be available on github repository

## References

[1] J. S. Brownstein, C. C. Freifeld, and L. C. Madoff, “Digital Disease Detection — Harnessing the Web for Public Health Surveillance,” The New England journal of medicine, vol.360, no. 21, pp. 2153–2157, May 2009. [Online]. Available: https://www.ncbi.nlm.nih.gov/pmc/articles/PMC2917042/

[2] G. Eysenbach, “Infodemiology: tracking flu-related searches on the web for syndromic surveillance.” AMIA… Annual Symposium proceedings / AMIA Symposium. AMIA Symposium, pp. 244–248, 2006.

[3] K. Espina and M. R. J. E. Estuar, “Infodemiology for Syndromic Surveillance of Dengue and Typhoid Fever in the Philippines,” Procedia Computer Science, vol. 121, pp. 554–561, Jan. 2017. [Online]. Available: http://www.sciencedirect.com/science/article/pii/S1877050917322731

[4] V. Gianfredi, N. L. Bragazzi, M. Mahamid, B. Bisharat, N. Mahroum, H. Amital, and M. Adawi, “Monitoring public interest toward pertussis outbreaks: an extensive Google Trends–based analysis,” Public Health, vol. 165, pp. 9–15, Dec. 2018. [Online]. Available: http://www.sciencedirect.com/science/article/pii/S0033350618302828

[5] G. Eysenbach, “Infodemiology and Infoveillance: Tracking Online Health Information and Cyberbehavior for Public Health,” American Journal of Preventive Medicine, vol. 40, no. 5, Supplement 2, pp. S154–S158, May 2011. [Online]. Available: http://www.sciencedirect.com/science/article/pii/S0749379711000882

[6] R. Ling and J. Lee, “Disease Monitoring and Health Campaign Evaluation Using Google Search Activities for HIV and AIDS, Stroke, Colorectal Cancer, and Marijuana Use in Canada: A Retrospective Observational Study,” JMIR Public Health Surveill, vol. 2, no. 2, 2016.

[7] S. Hamid, M. Y. Mir, and G. K. Rohela, “Novel coronavirus disease (COVID-19): a pandemic (epidemiology, pathogenesis and potential therapeutics),” New Microbes and New Infections, vol. 35, p. 100679, May 2020. [Online]. Available: http://www.sciencedirect.com/science/article/pii/S2052297520300317

[8] K. Chong Ng Kee Kwong, P. R. Mehta, G. Shukla, and A. R. Mehta, “COVID-19, SARS and MERS: A neurological perspective,” Journal of Clinical Neuroscience, May 2020. [Online]. Available: http://www.sciencedirect.com/science/article/pii/S0967586820311851

[9] S. E. Jordan, S. E. Hovet, I. C.-H. Fung, H. Liang, K.-W. Fu, and Z. T. H. Tse, “Using Twitter for Public Health Surveillance from Monitoring and Prediction to Public Response,” Data, vol. 4, no. 1, p. 6, Mar. 2019, number: 1 Publisher: Multidisciplinary Digital Publishing Institute. [Online]. Available: https://www.mdpi.com/2306-5729/4/1/6

[10] A. Abd-Alrazaq, D. Alhuwail, M. Househ, M. Hamdi, and Z. Shah, “Top Concerns of Tweeters During the COVID-19 Pandemic: Infoveillance Study,” Journal of medical Internet research, vol. 22, no. 4, p. e19016, 2020.

[11] L. Sinnenberg, A. M. Buttenheim, K. Padrez, C. Mancheno, L. Ungar, and R. M. Merchant, “Twitter as a tool for health research: a systematic review,” American journal of public health, vol. 107, no. 1, pp. e1–e8, 2017.

[12] J. Y. Breland, L. M. Quintiliani, K. L. Schneider, C. N. May, and S. Pagoto, “Social media as a tool to increase the impact of public health research,” American journal of public health, vol. 107, no. 12, p. 1890, 2017.

[13] A. J. Yepes, A. MacKinlay, and B. Han, “Investigating public health surveillance using twitter,” in Proceedings of BioNLP 15, 2015, pp. 164— 170.

[14] C. Chew and G. Eysenbach, “Pandemics in the age of twitter: content analysis of tweets during the 2009 h1n1 outbreak,” PloS one, vol. 5, no. 11, 2010.

[15] J. E. C. Saire, “Building intelligent indicators to detect dengue epidemics in brazil using social networks,” in 2019 IEEE Colombian Conference on Applications in Computational Intelligence (ColCACI). IEEE, 2019, pp. 1–5.

[16] J. E. C. Saire and R. C. Navarro, “What is the people posting about symptoms related to coronavirus in bogota, colombia?” arXiv preprint arXiv:2003.11159, 2020.

[17] J. E. Chire Saire, “Infoveillance based on social sensors to analyze the impact of covid19 in south american population,” 2020.

[18] J. E. Chire Saire and A. Pineda-Briseno, “Text mining approach to analyze coronavirus impact: Mexico city as case of study,” medRxiv, 2020. [Online]. Available: https://www.medrxiv.org/content/early/2020/05/12/2020.05.07.20094466

[19] S. Ear, “Towards effective emerging infectious diseases surveillance: Evidence from kenya, peru, thailand, and the u.s.-mexico,” 2012. [Online]. Available: https://siepr.stanford.edu/research/publications/towards-effective-emerging-infectious-diseases-surveillanceevidence-kenya-peru

